# Costs of digital adherence technologies for tuberculosis treatment support

**DOI:** 10.1101/2023.03.18.23287420

**Authors:** Ntwali Placide Nsengiyumva, Amera Khan, Maricelle Ma. Tarcela S. Gler, Mariceif Lopez, Danaida Marcelo, Mark C. Andrews, Karine Duverger, Shahriar Ahmed, Tasmia Ibrahim, Mona Lisa Morales, Andre Villanueva, Egwumo Efo, Baraka Onajare, Cristina Celan, Kevin Schwartzman

**Affiliations:** McGill International Tuberculosis Centre and Research Institute of the McGill University Health, Centre, Montréal, Québec, Canada; Stop TB Partnership, Geneva, Switzerland; DLSHSI, Makati City, Philippines; Health Through Walls, Haiti; icddr,b, Dhaka, Bangladesh; KNCV Manila, Philippines; KNCV Dar es Salaam, Tanzania; Center for Health Policies and Studies (PAS Center), Chisinau, Republic of Moldova

**Keywords:** Digital adherence technologies, cost, tuberculosis, pill sleeves, video-observed treatment

## Abstract

Digital adherence technologies (DATs) are increasingly used for tuberculosis (TB) adherence support, but information about their costs remains limited. We used microcosting to estimate health system costs (in 2022 US dollars) of 99DOTS pill sleeves and video-observed treatment (VOT), implemented in demonstration projects during 2018-21. Local cost estimates for standard directly observed treatment (DOT) were also obtained. The estimated per-person costs of 99DOTS for drug-sensitive (DS-) TB were $98, $106, and $174 in Bangladesh (n=719), the Philippines (n=396), and Tanzania (n=976) respectively. The estimated per-person costs of VOT were $1 154, $304, $457, and $661 in Haiti (n=87 DS-TB), Moldova (n=173 DS-TB), Moldova (n=135 drug-resistant [DR]-TB) and the Philippines (n=110 DR-TB) respectively. Health system costs of 99DOTS may be similar to or cheaper than standard DOT. VOT is considerably more expensive; labor cost offsets and/or economies of scale may yield savings relative to standard DOT in some settings.

**Summary:** In diverse settings, health system costs of 99DOTS pill sleeves may be similar to or cheaper than standard directly observed treatment for TB; video-observed treatment is considerably more expensive, but labor cost offsets and/or economies of scale may yield savings.

## Background

In the mission to cure and ultimately eliminate tuberculosis (TB), maintaining treatment adherence is a significant barrier (1). Individuals with TB disease must complete multidrug regimens typically lasting six or more months. Even small lapses in adherence can be associated with poorer treatment outcomes, including relapse with the potential for further transmission (2). TB prevention and care programs have often sought to improve adherence and hence treatment outcomes by using directly observed therapy (DOT) (3, 4). However, health system barriers, notably resource limitations, coupled with stigma, loss of autonomy and the heavy burden DOT clinic visits place on people with TB, can result in subpar outcomes, and adherence which may not exceed that with self-administered treatment (5-8). This has led the World Health Organization (WHO) to recommend community or home-based DOT over health facility-based DOT or unsupervised treatment (4).

The WHO defines a DOT provider as any person observing the individual with TB taking their medications in real time(4). By leveraging current advances in mobile technologies, person-centred treatment observation can be achieved by digital adherence technologies (DATs) such as medication sleeves, smart pill boxes, and video supported treatment. Moreover, real-time digital adherence information offers the possibility of tailoring treatment support to individual needs. However, before TB programs adopt these technologies as a central strategy for treatment support, evidence for their effectiveness must be robust. There is substantial evidence from demonstration projects highlighting feasibility and acceptability of DATs for TB treatment support; to date, evidence is more limited for clinical outcomes, when compared with other forms of treatment observation, or self-administered treatment (9-11). In principle, DATs can allow expansion of TB treatment supervision and support, while reducing burden on both persons with TB and their providers.

There has been limited information about the cost to TB programs of these technologies, and their real-world cost-effectiveness. Existing evidence comes largely from pilot and modeling studies (11, 12). We estimated the cost of two DATs currently recommended for use by the WHO (4), based on data from implementation studies in Bangladesh, Haiti, Moldova, the Philippines, and Tanzania.

## Methods

### Study design and tools

This multi-country cost analysis reflects DAT implementation projects funded by TB REACH Wave 6 (13). The goal was to estimate capital and recurrent costs of the DATs. We developed questionnaires and measurement tools to (1) describe the current standard of care (without DATs) for treatment of drug susceptible tuberculosis (DS-TB) and drug resistant tuberculosis (DR-TB); (2) document how the DATs used in each project were integrated into the local standard of care, and what changes in practice resulted from their introduction; and (3) capture all cost components of the DAT used during each project, including all related interventions and practice changes. Participating programs were provided with a series of cost analysis questionnaires, available online (14), and trained in their use via webinars. The present report is limited to the cost analysis and does not address cost-effectiveness. Data were collected prospectively, during the execution of each project.

### Study populations

Between 2018 and 2021, eligible adolescents and adults receiving DS-TB and DR-TB treatment were enrolled in DAT implementation projects in Bangladesh, Haiti, Moldova, the Philippines, and Tanzania. Key features of these projects and their participants are summarized in Table 1. They involved either the 99DOTS medication sleeves or video-observed treatment (VOT), also referred to as video-supported treatment (VST).

**Table 1.** Implementation projects and participants*

| **Country** | **DAT** | **Organization** | **Project description** | **Participants** |
| --- | --- | --- | --- | --- |
| Bangladesh | 99DOTS | icddr,b | Implementation of 99DOTS in private sector TB screening and treatment centres established by icddr,b under its social enterprise model in Dhaka. | 719 adults with DS-TB (mean age 34 (SD 27) were enrolled in the project which was conducted from April 2019 to July 2020. |
| Philippines | 99DOTS | KNCV | A project to assess 99DOTS use in the private sector in the Philippines, where data suggests 50% of patients in the country seek care. | 396 adults (≥15 years) with DS-TB were enrolled on 99DOTS at three private clinics based in metro Manila area from December 2018 to June 2020. |
| United Republic of Tanzania | 99DOTS | KNCV | The project involved mining communities in Tanzania in four rural districts using 99DOTS with SMS targeted educational messages and reminders. Patient dosing histories were used for counselling and for differentiated care (more intensive patient management), | 976 adult miners (>15 years) with DS-TB from 11 public health facilities were recruited at treatment initiation or during medication refill. 22 DOT nurses and 11 community health workers were engaged in the project between February 2019 and June 2020. |
| Haiti | VOT | Health through Walls, Inc. | A project using VOT to improve TB treatment adherence and outcomes for current and former prisoners in Haiti. | 87 incarcerated individuals with DS-TB were enrolled in the project between February 2019 to February 2020. The project used a commercial VOT platform. |
| Philippines | VOT | Hermano (San) Miguel Febres Cordero Medical Education Foundation, Inc | A pilot study to determine feasibility and acceptability of VOT in a high-burden, resource constrained DR-TB clinic in the Philippines where smartphones penetration is moderate and growing. | 110 adolescents and adults (>13 years) with DR-TB were enrolled to use VOT at six DR-TB clinics. Everyone received a smartphone with the VOT application pre-installed and with SIM cards. The study was conducted between December 2018 to June 2021. The project used a commercial VOT platform. |
| Republic of Moldova | VOT | Centre for Health Policies and Studies | A pilot project to scale up a locally developed VOT application | 173 adults with DS-TB and 135 adults with DR-TB were enrolled on a locally developed VOT platform from April 2020 to June 2021. |

\* Participants related to the costing exercise, the overall parent study might have had a higher number of participants CHW; community health worker. DAT; Digital adherence technology. DR-TB; drug resistant TB. DS-TB; drug susceptible TB. Icdrr,b; International Centre for Diarrhoeal Disease Research, Bangladesh. KNCV; Koninklijke Nederlandse Centrale Vereniging tot bestrijding der Tuberculose. TB; Tuberculosis. VOT; video-observed treatment; VST; video-supported treatment. SMS; short message service

### Cost analysis

We performed a combination of bottom-up and top-down microcosting for six TB-REACH DAT implementation projects. Seven other implementation projects pursued separate costing analyses, some of which have been published elsewhere e.g. the Ugandan project (15). Top-down costing used total amounts spent for DAT platform/infrastructure, systems/data management and technical support, and training activities. All other cost components were obtained using a bottom-up approach.

We conducted this analysis from the health system perspective. Hence costs borne by patients and their families were not tabulated. All costs were converted to 2022 US dollars using the respective countries’ inflation and exchange rates from the World Bank (16, 17). Costs reflected project expenses as reported by staff, and were grouped into two categories: fixed and variable costs. Variable costs included: 1) phones and accessories; 2) mobile data use; 3) adherence monitoring by health care workers (HCW) using the DAT platform; 4) HCW escalation/intervention in cases of non-adherence; 5) trainers; and 6) trainees. Two additional variable cost categories were specific to 99DOTS: 1) printing and shipping of medication sleeves, and 2) medication preparation (in case medication blister packs are not packaged within 99DOTS sleeves by the supplier).

Fixed costs included: 1) the platform and infrastructure, and 2) systems and data management/ technical support. Details of the costing tool are shown in the on-line appendix.

For each project, we estimated total costs and then prorated these per person treated for TB. As certain fixed costs (e.g., acquisition of the relevant platforms and computing infrastructure) can be substantial, we performed scenario analyses where the DAT was scaled up to support more persons during treatment. In these scenarios, variable costs remained unchanged, but we annuitized fixed technology costs based on a five-year lifespan and a 3% annual rate. We also explored scenarios where fixed technology/platform introduction and maintenance costs would be shared across expanded user numbers (i.e. 2X study population, 5X study population, 10X study population, and 100X study population), while maintaining the same variable costs as previously estimated.

To situate the DAT costs relative to the local standard of care; we evaluated incremental costs for DATs compared to in-person DOT, accounting for each study setting (i.e. duration of treatment and salary/wages of the individuals observing treatment). We supplemented this analysis by considering DOT costs from the existing literature. A threshold analysis was also performed, to capture the patient volumes at which DAT scale-up might become cost saving.

Each demonstration project participating in this study received local institutional ethics review board approval.

## Results

Between 2018 and 2021 the three 99DOTS projects enrolled a total of 2,091 patients: 719 in Bangladesh, 396 in the Philippines and 976 in Tanzania. During the same period, the three VOT projects enrolled a total of 505 patients: 87 in Haiti, 308 in Moldova and 110 in the Philippines. Further details are provided in Table 1. Adherence results are summarized in Table S5 of the online appendix. Total and per-person costs are provided in tables 2-5. Detailed component costs are listed in the appendix, tables S3 and S4.

**Table 2.** Total costs of 99DOTS during the implementation projects

| Cost in USD | Bangladesh | Philippines | Tanzania |
| --- | --- | --- | --- |
| Number of patients who used 99DOTS during the project | (n=719) | (n=396) | (n=976) |
| **Variable costs** | **\$43,718** | **\$24,854** | **\$100,860** |
| Phone and accessories | \$8,768 | \$296 | \$0 |
| Medication sleeves | \$4,993 | \$13,370 | \$9,748 |
| Medication preparation | \$1,137 | \$252 | \$426 |
| 99DOTS Calls/SMS costs | \$424 | \$573 | \$8,752 |
| Adherence monitoring by HCW using 99DOTS platform | \$17,547 | \$5,289 | \$1,064 |
| Escalation in case of non-adherence and HCW response | \$38 | \$439 | \$3,677 |
| DAT training for HCWs (Trainee costs) | \$4,727 | \$3,703 | \$47,784 |
| Additional training costs (Trainer costs) | \$6,084 | \$932 | \$29,410 |
| **Fixed costs** | **\$27,038** | **\$17,112** | **\$68,676** |
| Platform / infrastructure costs | \$14,772 | \$13,901 | \$10,509 |
| Systems, Data management and technical support | \$12,267 | \$3,211 | \$58,167 |
| **Total overall cost of 99DOTS** | **\$70,756** | **\$41,966** | **\$169,536** |

**Table 3.** Per person costs of 99DOTS during the implementation projects, without and with annuitization of capital costs

| Project site |  |  |  | With costs annuitized over the 5-year life span of servers and phones |  |  |
| --- | --- | --- | --- | --- | --- | --- |
| Country | Bangladesh | Philippines | Tanzania | Bangladesh | Philippines | Tanzania |
| Number of patients who used 99DOTS during the project | (n=719) | (n=396) | (n=976) | (n=719) | (n=396) | (n=976) |
| **Variable per patient cost** | **\$61** | **\$63** | **\$103** | **\$52** | **\$63** | **\$103** |
| Prorated per patient cost for phone and accessories * | \$12 | \$0.8 | \$0 | \$4 | \$0.8 | \$0 |
| Per patient cost for medication sleeves | \$7 | \$34 | \$10 | \$7 | \$34 | \$10 |
| Per patient medication preparation cost for entire duration of treatment | \$2 | \$0.6 | \$0.4 | \$2 | \$0.6 | \$0.4 |
| Total per patient cost of 99DOTS system call/SMS (TO AND FROM) for the entire duration of treatment | \$0.6 | \$2 | \$9 | \$0.6 | \$2 | \$9 |
| Per patient cost of 99DOTS adherence monitoring | \$24 | \$13 | \$1 | \$24 | \$13 | \$1 |
| Prorated per patient cost for escalation activity | \$0.1 | \$1 | \$4 | \$0.1 | \$1 | \$4 |
| Prorated per patient training cost for HCWs | \$7 | \$9 | \$49 | \$7 | \$9 | \$49 |
| Prorated per patient cost of running training sessions (not including time for HCW who are being trained) | \$9 | \$2 | \$30 | \$9 | \$2 | \$30 |
| **Fixed per patient cost\*#** | **\$38** | **\$43** | **\$70** | **\$28** | **\$16** | **\$60** |
| **Total per patient cost of 99DOTS** | **\$98** | **\$106** | **\$174** | **\$81** | **\$79** | **\$163** |

\* Equipment cost annuitized over a 5-year life span (phones, tablets and computers)

**Table 4.** Total costs of VOT at project sites

| Cost in USD | Haiti | Moldova<br>DS-TB | Moldova<br>DR-TB | Moldova<br>(All TB) | Philippines |
| --- | --- | --- | --- | --- | --- |
| Number of patients who used VOT during the project | (n=87) | (n=173) | (n=135) | (n=308) | (n=110) |
| **Variable cost** | **\$15,487** | **\$30,963** | **\$36,243** | **\$67,206** | **\$43,072** |
| Phone and accessories | \$9,960 | \$15,049 | \$17,615 | \$32,664 | \$15,636 |
| Data | \$732 | \$846 | \$991 | \$1,837 | \$0 |
| Adherence monitoring by HCW using VOT platform | \$2,845 | \$11,418 | \$13,365 | \$24,783 | \$14,601 |
| Escalation in case of non-adherence and HCW response | \$750 | \$284 | \$332 | \$616 | \$2,934 |
| DAT Training for HCWs (Trainee costs) | \$1,201 | \$3,366 | \$3,940 | \$7,306 | \$2,867 |
| Additional training costs (Trainer costs) | \$0 | \$0 | \$0 | \$0 | \$7,035 |
| **Fixed cost** | **\$84,949** | **\$21,714** | **\$25,416** | **\$47,130** | **\$29,584** |
| Platform / infrastructure | \$31,149 | \$18,429 | \$21,571 | \$40,000 | \$27,327 |
| Systems, Data management and technical support | \$53,800 | \$3,285 | \$3,845 | \$7,130 | \$2,257 |
| **Total overall cost of VOT** | **\$100,436** | **\$52,677** | **\$61,659** | **\$114,336** | **\$72,656** |

**Table 5.**
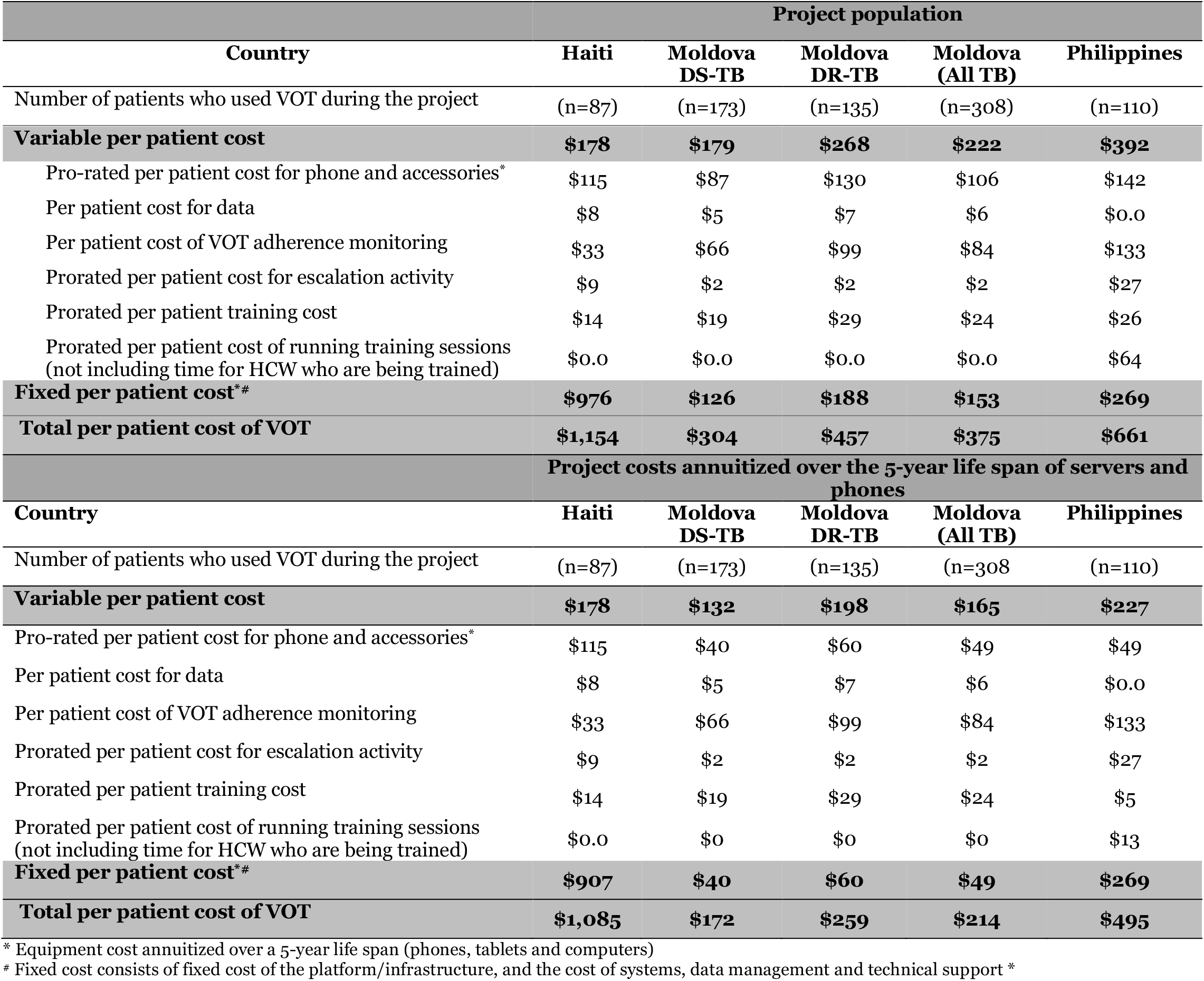
Per person VOT costs

| Project population |  |  |  |  |  |
| --- | --- | --- | --- | --- | --- |
| Country | Haiti | Moldova<br>DS-TB | Moldova<br>DR-TB | Moldova<br>(All TB) | Philippines |
| Number of patients who used VOT during the project | (n=87) | (n=173) | (n=135) | (n=308) | (n=110) |
| **Variable per patient cost** | **\$178** | **\$179** | **\$268** | **\$222** | **\$392** |
| Pro-rated per patient cost for phone and accessories* | \$115 | \$87 | \$130 | \$106 | \$142 |
| Per patient cost for data | \$8 | \$5 | \$7 | \$6 | \$0.0 |
| Per patient cost of VOT adherence monitoring | \$33 | \$66 | \$99 | \$84 | \$133 |
| Prorated per patient cost for escalation activity | \$9 | \$2 | \$2 | \$2 | \$27 |
| Prorated per patient training cost | \$14 | \$19 | \$29 | \$24 | \$26 |
| Prorated per patient cost of running training sessions (not including time for HCW who are being trained) | \$0.0 | \$0.0 | \$0.0 | \$0.0 | \$64 |
| **Fixed per patient cost\*#** | **\$976** | **\$126** | **\$188** | **\$153** | **\$269** |
| **Total per patient cost of VOT** | **\$1,154** | **\$304** | **\$457** | **\$375** | **\$661** |
| Project costs annuitized over the 5-year life span of servers and phones |  |  |  |  |  |
| Country | Haiti | Moldova<br>DS-TB | Moldova<br>DR-TB | Moldova<br>(All TB) | Philippines |
| Number of patients who used VOT during the project | (n=87) | (n=173) | (n=135) | (n=308) | (n=110) |
| **Variable per patient cost** | **\$178** | **\$132** | **\$198** | **\$165** | **\$227** |
| Pro-rated per patient cost for phone and accessories* | \$115 | \$40 | \$60 | \$49 | \$49 |
| Per patient cost for data | \$8 | \$5 | \$7 | \$6 | \$0.0 |
| Per patient cost of VOT adherence monitoring | \$33 | \$66 | \$99 | \$84 | \$133 |
| Prorated per patient cost for escalation activity | \$9 | \$2 | \$2 | \$2 | \$27 |
| Prorated per patient training cost | \$14 | \$19 | \$29 | \$24 | \$5 |
| Prorated per patient cost of running training sessions (not including time for HCW who are being trained) | \$0.0 | \$0 | \$0 | \$0 | \$13 |
| **Fixed per patient cost\*#** | **\$907** | **\$40** | **\$60** | **\$49** | **\$269** |
| **Total per patient cost of VOT** | **\$1,085** | **\$172** | **\$259** | **\$214** | **\$495** |

\* Equipment cost annuitized over a 5-year life span (phones, tablets and computers)

### 99DOTS

The estimated total cost of 99DOTS in the three implementation projects was $70 756, $41 966 and $169 633 overall, or $98, $106, and $174 per person treated for DS-TB in Bangladesh, the Philippines, and Tanzania respectively. Variable costs accounted for 62%, 59% and 59% of the total in Bangladesh, the Philippines, and Tanzania respectively. Tables 2 and 3 list sub-category costs and appendix table S3 lists detailed component costs.

The main cost drivers for 99DOTS varied across project sites. Key components included adherence monitoring by personnel (24% of costs) in Bangladesh, medication sleeves (32%) in the Philippines and training activities (43%) in Tanzania.

In scenario analyses we evaluated the variation in per-person costs as 99DOTS was potentially scaled to larger numbers of people treated for TB and their providers, while maintaining the same total fixed costs, and the same variable costs per patient. When the number of persons served by the platform was increased one hundred-fold, we estimated 47%, 41% and 40% decreases in cost per person to $55, $63, and $104 in Bangladesh, the Philippines, and Tanzania respectively (Figure 1). In all scenario analyses, the most influential cost components remained the same (as they all belonged to the variable cost category).

**Figure 1.**
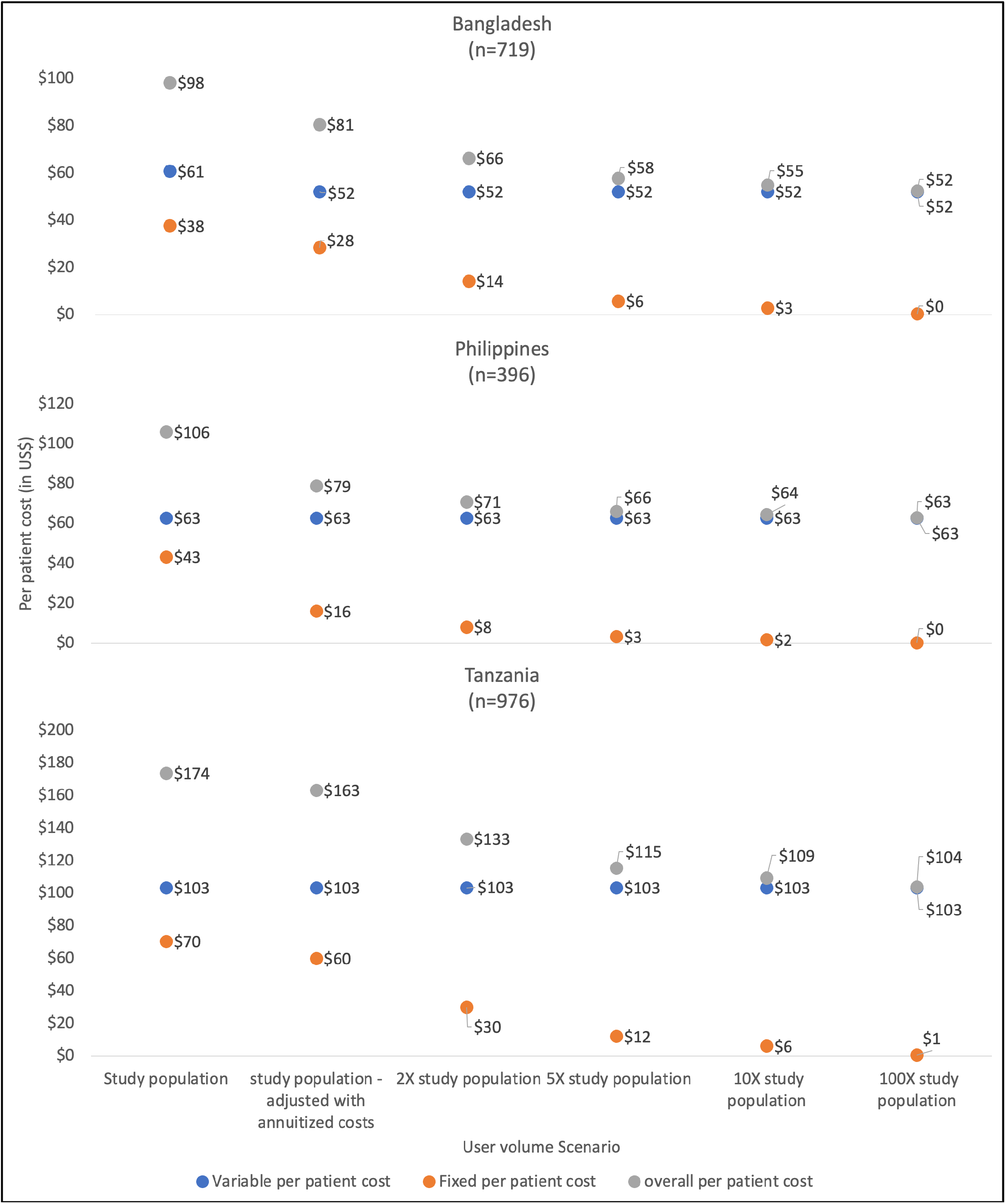
DOT scale-up scenario analysis scenarios where fixed technology/platform introduction and maintenance costs would be shared across expanded user numbers (i.e. 2X study population, 5X study population, 10X study population and 100X study population), while maintaining the same variable costs

### VOT

From the projects in Haiti (DS-TB), Moldova (all persons treated for TB), and the Philippines (DR-TB), estimated overall costs for VOT were $100 436, $114 336, and $72 656 respectively. Costs per person treated were $1 154, $375, and $661 respectively. Variable costs accounted for 15%, 59% and 59% of the total in Haiti (DS-TB), Moldova (all TB patients) and the Philippines (DR-TB) respectively. Tables 4 and 5 list sub-category costs, while detailed component costs are provided in appendix table S4.

The largest cost component in Haiti related to systems, data management and technical support which together accounted for 54% of the total. VOT platform and infrastructure accounted for 35% and 38% of the total cost in Moldova and the Philippines respectively.

In the scenario analysis, when the number of persons served by the same platform was increased one hundred-fold; we estimated an 83%, 56% and 54% decrease in cost, with per-person costs falling to $187, $165, and $229 in Haiti (DS-TB), Moldova (all TB patients), and the Philippines (DR-TB) respectively (figure 2). In that scenario, the largest cost component in Haiti (DS-TB) became the purchase of phones 61%, while adherence monitoring then accounted for 51% and 58% of the total cost in Moldova (all TB patients) and the Philippines respectively.

**Figure 2.**
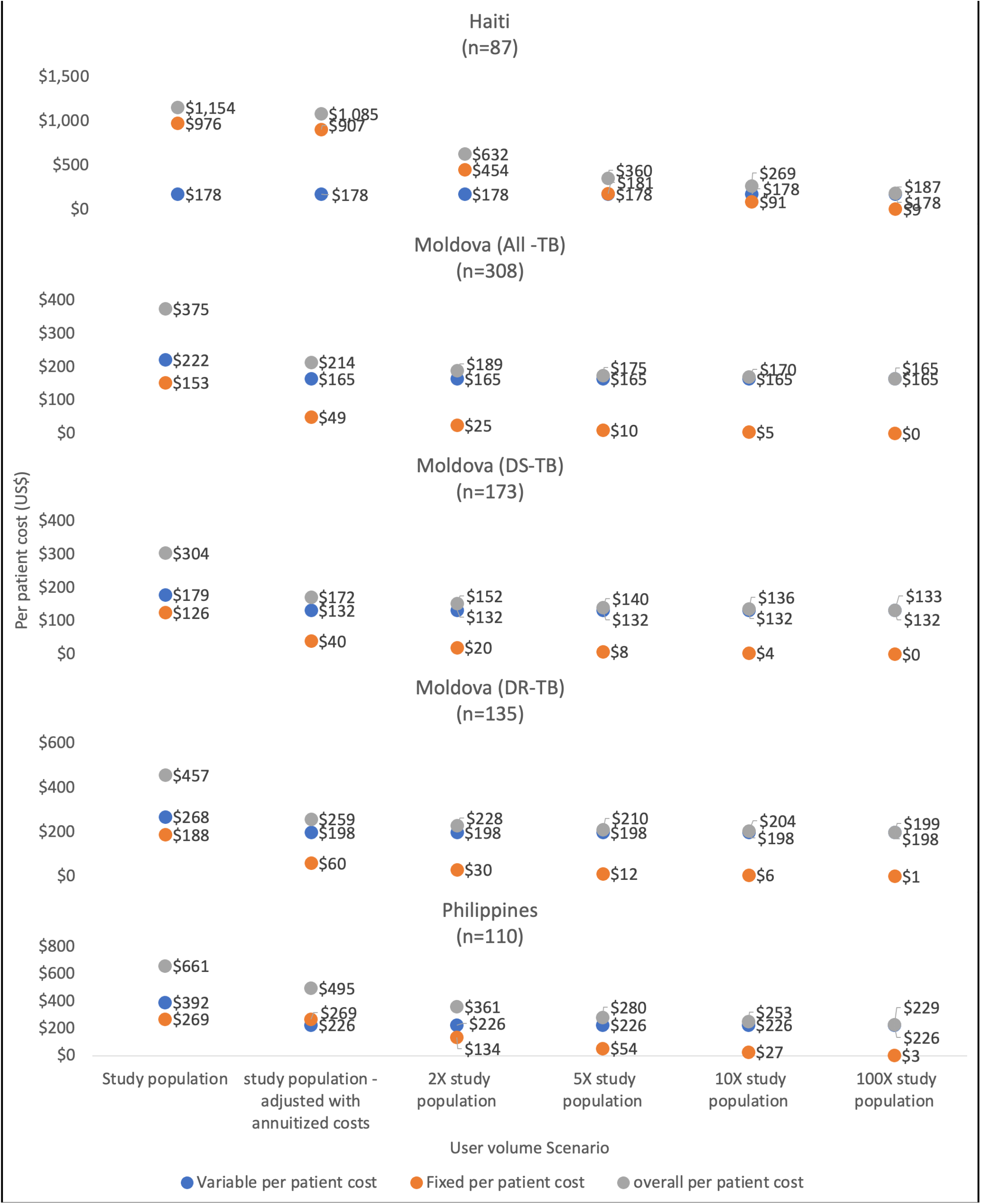
VOT scale-up scenario analysis scenarios where fixed technology/platform introduction and maintenance costs would be shared across expanded user numbers (i.e. 2X study population, 5X study population, 10X study population and 100X study population), while maintaining the same variable costs

### Cost of DAT vs DOT

We estimated costs for DAT-based treatment as compared to DOT in the different project settings, as detailed in table 6.

**Table 6:** DAT vs DOT costs ^*^

| 99DOTS sites |  |  |  | VOT sites |  |  |  |  |
| --- | --- | --- | --- | --- | --- | --- | --- | --- |
|  | Bangladesh | Philippines | Tanzania | Haiti | Moldova DS-TB | Moldova DR-TB | Moldova (All TB) | Philippines |
| **Crude estimate for DOT cost for the standard of care - derived from the costing tool** |  |  |  |  |  |  |  |  |
| Location of DOT - during the study | Health facility | Health facility | Home based | Prison | Health facility | Health facility | Health facility | Health facility or Home |
| Hourly wage of person offering DOT support | \$0.83 | \$1.40 | \$0.00 | \$4.60 | \$11.20 | \$11.20 | \$11.20 | \$2.88 |
| Duration of DOT visit - in minutes | 45 | 45 | 0 | 45 | 15 | 15 | 15 | 2 |
| **Total per patient cost** |  |  |  |  |  |  |  |  |
| DOT cost from study data | \$74 | \$176 | \$0 | \$414 | \$336 | \$504 | \$427 | \$17 |
| DOT cost from the literature | \$36 (18) | \$155 (19) | \$89 (21) | \$49 (20) | | | | \$166(19) |
| Annuitized cost of DAT | \$81 | \$79 | \$178 | \$1,085 | \$172 | \$259 | \$214 | \$495 |
| **Incremental cost for the DAT** |  |  |  |  |  |  |  |  |
| Incremental cost of DAT vs standard of care (**Study data as comparator**) * | \$7 | -\$97 | \$178 | \$671 | -\$164 | -\$245 | -\$213 | \$478 |
| Incremental cost of DAT vs standard of care (**Prior published data as comparator**) # | \$45 | -\$76 | \$89 | \$1,036 | | | | \$329 |

\* Negative incremental costs indicate savings for the DAT relative to standard of care

#### 99DOT vs DOT

The 99DOTS projects were conducted in settings where TB treatment is ordinarily observed by either a health facility worker, community health care worker or family members. Program personnel estimated that 30%, 20% and 0% of patients would remain on traditional DOT after the introduction of 99DOTS in Bangladesh, Philippines, and Tanzania, respectively. Such individuals included those receiving TB retreatment, persons without access to a mobile phone, persons residing outside the clinic’s catchment area, hospitalised patients, persons with extrapulmonary TB on ≥12 months treatment regimens, and persons unwilling to provide consent. From the Tanzania health system perspective, DOT itself does not imply health system costs, and hence DOT costs to the health system are not offset by use of 99DOTS, since treatment support is ordinarily provided (unpaid) by family members.

In a threshold analysis, we explored the patient volumes required for 99DOTS to be cost saving when compared to DOT. In Bangladesh, when we used our own DOT cost estimates, 99DOTS was associated with a $7 incremental cost with the patient numbers enrolled. When we used DOT costs published elsewhere the incremental cost was $45 *(18)*. We estimated that an increase of >30% in patient volumes from the study population would render 99DOTS cost-saving. In the Philippines, 99DOTS was cost saving with existing patient volumes, using both our own DOT cost estimates and those published elsewhere *(19)*. In Tanzania, there was no possibility of health system cost savings with 99DOTS compared to DOT, as the latter relied on family members at no cost to the health care system.

#### VOT vs DOT

Program personnel estimated that 15%, 10% and 50% of persons treated for TB would remain on traditional DOT after the implementation of VOT in Haiti, Moldova, and the Philippines, respectively. Use of DAT in Haiti was associated with $671 and $1,036 incremental costs when compared to study DOT cost and published literature DOT cost respectively *(20)*. In a threshold analysis, we estimated that a four-fold increase in the number of patients covered in Haiti would be cost saving. In Moldova, VOT was cost saving with existing patient volumes, using our own DOT cost estimates. 69% and 51% of the DS-TB and DR-TB study populations actually enrolled respectively, would have been sufficient to generate cost savings in Moldova. In the Philippines, VOT costs per person exceeded those for DOT even with all fixed costs excluded, meaning that expanding the number of patients covered by VOT (i.e. economies of scale) could not result in cost savings.

## Discussion

This cost analysis for two digital adherence technologies covered a wide range of settings with diverse populations, socioeconomic conditions, and TB epidemiology. Implementation costs were substantial when limited to a small number of persons with TB—particularly infrastructure and training costs. However, if the DAT programs are scaled up to cover larger numbers of persons with TB, the health system cost per treatment course would fall and could in fact become cheaper than paid in-person DOT.

The same infrastructure can only be stretched so far; a better understanding of its capacity will be essential in understanding cost and budgetary impact of DAT expansion. Moving forward, it will also be important to account for potential cost savings to the health system, to the extent that in-person DOT is reduced), and especially for potential cost savings to people and families affected by TB.

This analysis had several strengths, including a diversity of real-world settings reflective of those where TB care is provided. It also used carefully gathered microcosting data. We explicitly considered scenarios where user volume could be expanded, in order to better harness the necessary technical infrastructure.

We did not evaluate the effectiveness of the DATs. This has been addressed in other related publications and an analysis of the feasibility and acceptability of DATs in the TB REACH projects is forthcoming (22). The technologies themselves have recognized limitations: for example, messages received or not received by the 99DOTS platform do not necessarily equate with medication ingestion, or lack thereof (23, 24). Hence direct comparisons of DAT and DOT costs are only appropriate to the extent that clinical outcomes with the DAT in question are similar to or better than standard care; this point was explicitly not addressed in the current analysis. Importantly, we did not evaluate costs or savings for patients and family members, e.g. related to missed work hours or child care needs, which may be mitigated when in-person DOT is replaced by digital treatment support. Other studies have highlighted the importance of such savings in the DAT context (25, 26).

The high costs of DATs, especially VOT, are driven in large part by one-time infrastructure costs such as computing equipment and phones, initial configuration, and software licencing (see appendix table S3). For Haiti and Moldova where the cost of purchasing phones accounted for a significant portion of the per patient cost, the loan of phones to patients or their use of personal phones would drastically lower the cost of implementing VOT. The per patient cost was particularly higher for Haiti because of higher cost for hardware at the beginning of project as there was a learning curve when evaluating whether tablets or phones worked best for intervention. Asynchronous VOT offsets some recurrent costs associated with synchronous VOT, and improves flexibility by allowing persons with TB to record medication ingestion within an agreed range of time, even if they do not have internet access at that moment (27). Similarly, the use of compressed video files can lower data use costs. Less worker time is needed if the recordings are reviewed using higher playback speed, and/or computer-assisted recognition of pill swallowing (28, 29). However, this may not be sufficient to make VOT easily accessible to TB programs in low- and lower middle-income countries.,

Global DAT initiatives for TB are addressing the infrastructure cost burden by improving market access, procurement mechanisms, and supply chains (30). Our study complements this work by carefully documenting capital and operating expenditures, allowing for better planning and decisions by TB treatment programs (31). Our cost estimates for 99DOTS are similar to those from other TB programs which used this technology (15, 32). A recent study from the USA estimated that VOT was cheaper than in-person DOT provided by health care staff (25), for both the health care system and for persons with TB and their families. To the extent that DATs offload health care workers from in-person observation, this will reduce their net cost. However, there are also local factors which further shape overall costs such as the cost of internet and SMS, the DAT and platform used, specific infrastructure used and patient population served, labor costs and treatment duration. There remain important barriers beyond widespread internet connectivity and infrastructure: these include limited availability, accessibility, and affordability of some technologies for persons in resource-limited areas, and similarly the availability and affordability of technical personnel needed to support the TB clinics (22, 33). Hence real-world cost, effectiveness, and implementation data from high TB-incidence, lower-income settings will remain paramount.

## Conclusion

Advances in the usability and acceptability of digital adherence technologies, coupled with widespread internet access and mobile phone use, make them viable tools for person-centred adherence support. However, economic evaluations are limited to date. Our analysis suggests that 99DOTS may be affordable to TB programs in diverse settings, particularly if used at scale. VOT appears less affordable for lower-income countries at present, although costs for both technologies will be reduced if the same infrastructure and hardware can support more patients.

## Data Availability

All data produced in the present study are available upon reasonable request to the authors

