## Supplemental tables for "Costs of digital adherence technologies for tuberculosis treatment support"

DAT Supplemental Appendix

#### Table S1. Costing tool - 99DOTS

| Cost components | Details/Formula * |
|---|---|
| Project and respondent information | |
| 1. Name of the project | Text |
| 2. Country | Text |
| 3. Name of respondent | Text |
| 4. Position of respondent | Text |
| 5. e-mail of respondent | Text |
| 6. Phone | Text |
| 7. Local currency used in the country | Text |
| 8. Date the questionnaire was filled | YYYY - MM - DD |
| 9. Was anything donated to the project? (e.g. equipment, technical support, training sessions offered as donations, etc.) | Yes, No |
| 10. List detailed items and services that were donated to the project | Text |
| Population | |
| I. Number of patients who used 99DOTS during the project | N |
| a) Phone and accessories | For patients |
| A1. Number of phones that were purchased | N |
| A2. Unit cost of phone purchased | $ |
| A3. Number of SIM cards that were purchased | N |
| A4. Unit cost of SIM card | $ |
| A5. Number of charger or solar batteries that were purchased | N |
| A6. Unit cost of charger or solar batteries | $ |
| A7. Number of phone lines provided with airtime | N |
| A8. Total cost for airtime per phone line for the entire duration of the project | $ |
| A9. Total cost of phones (includes phone, SIM card and airtime) | (A1*A2) + (A3*A4) + (A5*A6) + (A7*A8) |
| Pro-rated per patient cost for phone and accessories | A9/I |
| b) Platform / infrastructure costs | |
| B1. Initial configuration/customization costs (setting up phonelines and or platform at sites) | $ |
| B2. Type of configuration (portable, toll-free or SMS) | Text |
| B3. Total fixed cost of renting toll free phone lines / SMS line or any other system used to support 99DOTS platform for the entire duration of the project | $ |
| Per patient fixed cost of the platform/infrastructure | (B1 + B3)/I |
| c) Envelopes | |
| C1. Total cost of 99DOTS envelope printing (including secondary packaging, labels, etc.) | $ |
| C2. Total cost for shipping of 99DOTS envelopes (including customs, etc.) | $ |
| Per patient cost for envelopes | (C1+C2)/I |
| d) Medication Preparation | |
| D1. Typical amount of time to wrap and prepare one medication envelope (in minutes) | Minutes |
| D2. Type of staff wrapping medication | Job Category |
| D3. Wage of staff (per hour) | $ |
| D4. Cost of labour for wrapping per envelope (Total time for wrapping an envelope) | D1*D3/60 |
| D5. Number of envelopes required per patient for the entire duration of treatment | N |
| Per patient preparation cost for entire duration of treatment | D4*D5 |
| e) 99DOTS Calls/SMS costs | |
| E1. Average number of calls/SMS made by a patient to 99DOTS during the entire duration of treatment | N |
| E2. Amount the project paid per call/SMS made by a patient to 99DOTS | $ |
| E3. Per patient call/SMS to 99DOTS cost for entire duration of treatment | E1*E2 |
| E4. Average number (per patient) of 99DOTS reminder/education SMS sent by the system for missed doses | N |
| E5. cost the project has to pay per 99DOTS system SMS | $ |
| E6. per patient cost of 99DOTS adherence reminders for the entire duration of treatment | E4*E5 |
| Total per patient cost (TO AND FROM) 99DOTS system call/SMS for entire duration of treatment | E3 + E6 |
| f) Adherence monitoring by HCW using 99DOTS platform | |
| F1. Type of HCW who does adherence monitoring using 99DOTS platform | Job Category |
| F2. HCW wage (per hour) | $ |

| Cost components | Details/Formula * |
|---|---|
| F3. Typical amount of time (in min) spent by HCW monitoring adherence on the platform per patient (duration of project) | N |
| Per patient cost of 99DOTS adherence monitoring | F3*F2/60 |
| g) Systems, Data management and technical support | |
| G1. Amount spent for staff working on data management for 99DOTS platform (salaries paid for support of DAT platform) | $ |
| G2. Number of days that technical support was provided, or number of times specific technical services were provided (beyond initial setup) | N |
| G3. Cost per day or per activity reported in G2 | $ |
| G4. Total cost of technical support for 99DOTS platform | G2*G3 |
| G5. Amount paid for health facility hardware (laptops, desktop, or tablet computer to track adherence) | $ |
| G6. Payments made for monthly plans for technical services such as data plans and phone plans for HCW and health facility | $ |
| Per patient cost for Systems, Data management and technical support | (G1 + G4 + G5 + G6)/I |
| h) Escalation in case of non-adherence and HCW response | (improve adherence) |
| Escalation related phone calls: | |
| H1. Number of patients who required follow up by phone as part of adherence escalation procedure | N |
| H2. Average number of phone calls made by HCW per patient (for entire duration of the project) | N |
| H3. Average HCW time per call (in minutes) required to speak to a patient who requires adherence follow up | minutes |
| H4. Total time (in minutes) spent by HCW with patients on a phone for adherence follow up | H1*H2*H3 |
| H5. Type of HCW who usually makes phone calls | Job Category |
| H6. Average HCW wage (per hour) | $ |
| H7. Additional fees associated with phone call follow up (per call) | $ |
| H8. Total cost for Phone calls | (H4*H6/60min) + (H1*H2*H7) |
| Escalation related Home Visits: | |
| H9. Number of patients who required a home visit as part of escalation procedure | N |
| H10.Average number of home visits per individual who requires home visits | N |
| H11.Amount of time per visit (including travel) | minutes |
| H12. Total time (in minutes) spent by HCW on home visits for patients who required adherence follow up | H9*H10*H11 |
| H13.Type of HCW doing home visit | Job Category |
| H14.HCW wage (per hour) | $ |
| H15.Additional costs for HCW per visit (e.g., Travel/transport costs, incentives, etc.) | $ |
| H16. Total cost for home visits | (H12*H14/60) + (H9*H10*H15) |
| Pro-rated per patient cost for escalation activity | (H8 + H16)/I |
| i) DAT Training for HCWs *(Trainee costs)* | List up to three |
| 1st Category of HCW trained | Job Category 1 |
| I1.1 Number of Category 1 trained | N |
| I1.2 Typical amount of time for training (in minutes) (per HCW in category 1) | Minutes |
| I1.3 Total time in minutes for training all HCW in category 1 | I1.1*I1.2 |
| I1.4 HCW wage (per hour) | $ |
| I1.5 Total cost for training all HCW in category 1 | I1.3*I1.4/60 |
| 2nd Category of HCW trained | Job Category 2 |
| I2.1 Number of Category 2 trained | N |
| I2.2 Typical amount of time for training (in minutes) (per HCW in category 2) | Minutes |
| I2.3 Total time in minutes for training all HCW in category 2 | I2.1*I2.2 |
| I2.4 HCW wage (per hour) (category 2) | $ |
| I2.5 Total cost for training all HCW in category 2 | I2.3*I2.4/60 |
| 3rd Category of HCW trained | Job Category 3 |
| I3.1 Number of Category 3 trained | N |
| I3.2 Typical amount of time for training (in minutes) (per HCW in category 3) | Minutes |
| I3.3 Total time in minutes for training all HCW in category 3 | I3.1*I3.2 |
| I3.4 HCW wage (per hour) (category 3) | $ |
| I3.5 Total cost for training all HCW in category 3 | I3.3*I3.4/60 |
| I4. Total cost for training | I1.5 + I2.5 +I3.5 |
| Pro rated per patient training cost | I4/III |
| j) Additional training costs *(Trainer costs)* | |
| Choose the training scenario for your project | |
| Scenario I: training was provided by individuals who were paid for each training session | |

| Cost components | Details/Formula * |
|---|---|
| J1. Most common job category of staff providing the training sessions for HCW on how to use the 99DOTS | Job category (trainer) |
| J2. Typical amount of time spent by a trainer on training (including preparation and delivery) (in hours) | Hours |
| J3. Approximate hourly salary of someone doing training (trainer) | $ |
| J4. Number of trainers | N |
| J5. Total trainers' wage directly related to 99DOTS training | J2*J3*J4 |
| Scenario II: training was conducted by an outside organization or paid as a package | |
| J6. Total trainers' cost | $ |
| J7. Additional expenditures related to training (including travel, training venue, subsistence for trainers and trainees, etc. but not time spent by, or salary paid to trainers or trainees)? Specify in column C and enter the cost in column C | $ |
| Pro rated per patient cost of running training sessions (not including time for HCW who are being trained) | IF Scenario I: (J5 + J7)/III<br>IF Scenario II: (J6 + J7)/III |

| | |
|---|---|
| **Total per patient cost for 99DOTS is the SUM of the following categories** | **Pro-rated per patient cost for phone and accessories** |
| | **Per patient fixed cost of the platform/infrastructure** |
| | **Per patient cost for envelopes** |
| | **Per patient preparation cost for entire duration of treatment** |
| | **Total per patient cost (TO AND FROM) 99DOTS system call/SMS for entire duration of treatment** |
| | **Per patient cost of 99DOTS adherence monitoring** |
| | **Per patient cost for Systems, Data management and technical support** |
| | **Pro-rated per patient cost for escalation activity** |
| | **Pro-rated per patient training cost** |
| | **Pro-rated per patient cost of running training sessions (not including time for HCW who are being trained)** |

N=Numeric,
* text in blue Indicates calculated fields

#### Table s2. Costing tool - VOT

| Cost components | Details/Formula * |
|---|---|
| **Project and respondent information** | |
| 1. Name of the project | Text |
| 2. Country | Text |
| 3. Name of respondent | Text |
| 4. Position of respondent | Text |
| 5. e-mail of respondent | Text |
| 6. Phone | Text |
| 7. Local currency used in the country | Text |
| 8. Date the questionnaire was filled | YYYY - MM - DD |
| 9. Was anything donated to the project? (e.g. equipment, technical support, training sessions offered as donations,etc.) | Yes, No |
| 10. List detailed items and services that were donated to the project | Text |
| **Population** | |
| I. Type of patients (DS-TB or DR-TB) | Text |
| II. Length of a full course of treatment (in months) | N |
| III. Number of patients who were followed using MERM (evriMED or FLOW e-monitors) during the project | N |
| a) Phone and accessories | |
| A1. Number of phones that were purchased | N |
| A2. Unit cost of phone purchased | $ |
| A3. Number of SIM cards that were purchased | N |
| A4. Unit cost of SIM card | $ |
| A5. Number of charger or solar batteries that were purchased | N |
| A6. Unit cost of charger or solar batteries | $ |
| A7. Number of phone lines provided with airtime | N |
| A8. Total cost for airtime per phone line for the entire duration of the project | $ |
| A9. Total cost of phones (includes phone, SIM card and airtime) | (A1*A2) + (A3*A4) + (A5*A6) + (A7*A8) |
| Pro rated per patient cost for phone and accessories | A9/III |
| **b) Data** | |
| B1. Average size of a video recording in Megabytes (MB) | N |
| B2. Cost per MB of data | $ |
| B3. Total number of video calls/recordings made by all patients in the project | N |
| B4. Total cost for data for all patients | B1*B2*B3 |
| Per patient cost for data | B4/III |
| **c) Platform / infrastructure costs** | |
| C1. Total amount spent on software licence (if applicable) for the entire duration of the project | $ |
| C2. Initial Configuration costs (setting up VDOT platform in country) | $ |
| C3. Total amount spent to develop a VDOT software or platform for a project not using an already existing system | $ |
| Per patient fixed cost of the platform/infrastructure | (C1 + C2 + C3)/III |
| **d) Adherence monitoring by HCW using VDOT platform** | |
| Routine adherence monitoring using VDOT | |
| D1. Type of HCW who does adherence monitoring using VDOT platform | Text |
| D2. HCW wage (per hour) | $ |
| D3. Typical amount of time (in min) spent by HCW monitoring adherence on the platform per patient (duration of project) | N |
| D4. Per patient cost of VDOT routine adherence monitoring | D3*D2/60 |
| Video recordings viewing | |
| D5. Type of HCW who usually views video recordings | Text |
| D6. HCW wage (per hour) | $ |
| D7. Average number of minutes per patient per recording | N |
| D8. Average number of recordings made by a patient in the project | B3/III |
| D9. Per patient average amount of time (in minutes) of recordings over the course of treatment | D7*D8 |
| D10. Per patient cost of viewing video recordings | D9*D6/60 |

| Cost components | Details/Formula * |
|---|---|
| Per patient cost of VDOT adherence monitoring (routine + viewing of recordings) | D4 + D10 |
| e) Systems, Data management and technical support | |
| E1. Amount spent for staff working on data management for VDOT platform (salaries paid for support of DAT platform) | $ |
| E2. Number of days that technical support was provided or number of times specific technical services were provided (beyond initial setup) | N |
| E3. Cost per day or per activity reported in E2 | $ |
| E4. Total cost of technical support for VDOT platform | E2*E3 |
| E5. Amount paid for health facility hardware (laptops, desktop or tablet computer to track adherence) | $ |
| E6. Payments made for monthly plans for technical services such as data plans and phone plans for HCW and health facility | $ |
| Per patient cost for Systems, Data management and technical support | (E1 + E4 + E5 + E6)/III |
| f) Escalation in case of non-adherence and HCW response | |
| Escalation related phone calls: | |
| F1. Number of patients who required follow up by phone as part of adherence escalation procedure | N |
| F2. Average number of phone calls made by HCW per patient (for entire duration of the project) | N |
| F3. Average HCW time per call (in minutes) required to speak to a patient who requires adherence follow up | minutes |
| F4. Total time ( in minutes) spent by HCW with patients on a phone for adherence follow up | F1*F2*F3 |
| F5. Type of HCW who usually makes phone calls | Job Category |
| F6. Average HCW wage (per hour) | $ |
| F7. Additional fees associated with phone call follow up (per call) | $ |
| F8. Total cost for Phone calls | (F4*F6/60min) + (F1*F2*F7) |
| Escalation related Home Visits: | |
| F9. Number of patients who required a home visit as part of escalation procedure | N |
| F10.Average number of home visits per individual who requires home visits | N |
| F11.Amount of time per visit (including travel) | minutes |
| F12. Total time ( in minutes) spent by HCW on home visits for patients who required adherence follow up | F9*F10*F11 |
| F13.Type of HCW doing home visit | Job Category |
| F14.HCW wage (per hour) | $ |
| F15.Additional costs for HCW per visit (e.g. Travel/transport costs, incentives, etc.) | $ |
| F16. Total cost for Home visits | (F12*F14/60) + (F9*F10*F15) |
| Pro rated per patient cost for escalation activity | (F8 + F16)/III |
| g) DAT Training for HCWs (Trainee costs) | |
| 1st Category of HCW trained | Job Category 1 |
| G1.1 Number of HCW trained | N |
| G1.2 Typical amount of time for training (in minutes) (per HCW in category 1) | Minutes |
| G1.3 Total time in minutes for training all HCW in category 1 | G1.1*G1.2 |
| G1.4 HCW wage (per hour) | $ |
| G1.5 Total cost for training all HCW in category 1 | G1.3*G1.4/60 |
| 2nd Category of HCW trained | Job Category 2 |
| G2.1 Number of HCW trained | N |
| G2.2 Typical amount of time for training (in minutes) (per HCW in category 2) | Minutes |
| G2.3 Total time in minutes for training all HCW in category 2 | G2.1*G2.2 |
| G2.4 HCW wage (per hour) (category 2) | $ |
| G2.5 Total cost for training all HCW in category 2 | G2.3*G2.4/60 |
| 3rd Category of HCW trained | Job Category 3 |
| G3.1 Number of HCW trained | N |
| G3.2 Typical amount of time for training (in minutes) (per HCW in category 3) | Minutes |
| G3.3 Total time in minutes for training all HCW in category 3 | G3.1*G3.2 |
| G3.4 HCW wage (per hour) (category 3) | $ |
| G3.5 Total cost for training all HCW in category 3 | G3.3*G3.4/60 |
| G4. Total cost for training | G1.5 + G2.5 +G3.5 |
| Pro rated per patient training cost | G4/III |

| Cost components | Details/Formula * |
|---|---|
| h) Additional training costs (Trainer costs) | |
| Choose the training scenario for your project | |
| Scenario I: training was provided by individuals who were paid for each training session | |
| H1. Most common job category of staff providing the training sessions for HCW on how to use the DAT | Job category (trainer) |
| H2. Typical amount of time spent by a trainer on training (including preparation and delivery) (in hours) | Hours |
| H3. Approximate hourly salary of someone doing training (trainer) | $ |
| H4. Number of trainers | N |
| H5. Total trainers' wage directly related to VDOT training | H2*H3*H4 |
| Scenario II: training was conducted by an outside organization or paid as a package | |
| H6. Total trainers' cost | $ |
| H7. Additional expenditures related to training (including travel, training venue, subsistence for trainers and trainees, etc. but not time spent by or salary paid to trainers or trainees)? Specify in column C and enter the cost in column C | $ |
| Pro rated per patient cost of running training sessions (not including time for HCW who are being trained) | IF Scenario I: (H5 + H7)/III<br>IF Scenario II: (H6+H7)/III |

| | |
|---|---|
| **Total per patient cost for VDOT is SUM of the following categories** | **Pro-rated per patient cost for smart phone** |
| | **Per patient cost for data** |
| | **Per patient fixed cost of the platform/infrastructure** |
| | **Per patient cost of VDOT adherence monitoring by HCW** |
| | **Per patient cost for Systems, Data management and technical support** |
| | **Pro-rated per patient cost for escalation activity** |
| | **Pro-rated per patient cost for HCW training** |
| | **Pro-rated per patient trainers' cost** |

N=Numeric,

* Text in blue Indicates calculated fields

Table S3. Cost components for 99DOTS across the three study populations

| Cost components | Bangladesh | The Philippines | Tanzania |
|---|---|---|---|
| Population | | | |
| I. Number of patients who used 99DOTS during the project | 719 | 396 | 976 |
| a) Phone and accessories | | | |
| A1. Number of phones that were purchased | 22 | 0 | 0 |
| A2. Unit cost of phone purchased | $354.58 | $0 | $0 |
| A3. Number of SIM cards that were purchased | 22 | 296 | 0 |
| A4. Unit cost of SIM card | $11.79 | $1.00 | - |
| A5. Number of charger or solar batteries that were purchased | 0 | 0 | 0 |
| A6. Unit cost of charger or solar batteries | $0.00 | - | - |
| A7. Number of phone lines provided with airtime | 10 | 0 | 0 |
| A8. Total cost for airtime per phone line for the entire duration of the project | $70.74 | - | - |
| A9. Total cost of phones (includes phone, SIM card and airtime) | $8,767.63 | $296.00 | - |
| **Pro-rated per patient cost for phone and accessories** | **$12.19** | **$0.75** | **-** |
| b) Platform / infrastructure costs | | | |
| B1. Initial configuration/customization costs (setting up phonelines and or platform at sites) | $ 10,304.5 | $ 13,000.0 | $8000 |
| B2. Type of configuration (portable, toll-free or SMS) | - | toll-free SMS | $8,736.3 |
| B3. Total fixed cost of renting toll free phone lines / SMS line or any other system used to support 99DOTS | $4,467.2 | $901.3 | $1,772.8 |
| **Per patient fixed cost of the platform/infrastructure** | **$20.54** | **$35.10** | **$10.77** |
| c) Envelopes | | | |
| C1. Total cost of 99DOTS envelope printing (including secondary packaging, labels, etc.) | $4,922.33 | $13,000 | $3,628 |
| C2. Total cost for shipping of 99DOTS envelopes (including customs, etc.) | $70.74 | $370.00 | $6,120.00 |
| **Per patient cost for envelopes** | **$ 6.94** | **$33.76** | **$9.99** |
| d) Medication Preparation | | | |
| D1. Typical amount of time to wrap and prepare one medication envelope (in minutes) | 2 | 1 | 1 |
| D2. Type of staff wrapping medication | Research | TB nurse | Nurse |
| D3. Wage of staff (per hour) | $1.36 | $1.59 | $1.09 |
| D4. Cost of labour for wrapping per envelope (Total time for wrapping an envelope) | $0.05 | $0.03 | $0.02 |
| D5. Number of envelopes required per patient for the entire duration of treatment | 35 | 24 | 24 |
| **Per patient preparation cost for entire duration of treatment** | **$1.58** | **$0.64** | **$0.44** |
| e) 99DOTS Calls/SMS costs | | | |
| E1. Average number of calls/SMS made by a patient to 99DOTS during the entire duration of treatment | 180 | 128 | 99 |
| E2. Amount the project paid per call/SMS made by a patient to 99DOTS | $0.00 | $0.01 | $0.03 |
| E3. Per patient call/SMS to 99DOTS cost for entire duration of treatment | - | $1.02 | $3.27 |
| E4. Average number (per patient) of 99DOTS reminder/education SMS sent by the system for missed doses | 50 | 53 | 190 |
| E5. cost the project has to pay per 99DOTS system SMS | $0.01 | $0.01 | $0.03 |
| E6. per patient cost of 99DOTS adherence reminders for the entire duration of treatment | $0.59 | $0.42 | $5.70 |
| **Total per patient cost (TO AND FROM) 99DOTS system call/SMS for entire duration of treatment** | **$0.59** | **$1.45** | **$8.97** |
| f) Adherence monitoring by HCW using 99DOTS platform | | | |
| F1. Type of HCW who does adherence monitoring using 99DOTS platform | Research | TB nurse | Nurse |
| F2. HCW wage (per hour) | $1.36 | $1.59 | $1.09 |
| F3. Typical amount of time (in min) spent by HCW monitoring adherence on the platform per patient | 1080 | 504 | 60 |
| **Per patient cost of 99DOTS adherence monitoring** | **$24.41** | **$13.36** | **$1.09** |
| g) Systems, Data management and technical support | | | |
| G1. Amount spent for staff working on data management for 99DOTS platform (support of DAT platform) | $1,459.01 | $1,639.21 | $38,137.50 |
| G2. Number of days that technical support was provided, or times specific technical services provided | 35 | 15 | 6 |
| G3. Cost per day or per activity reported in G2 | $56.14 | $74.50 | $2,652.42 |
| G4. Total cost of technical support for 99DOTS platform | $1,965.04 | $1,117.50 | $15,914.52 |
| G5. Amount paid for health facility hardware (laptops, desktop, or tablet computer to track adherence) | $8,253.00 | $449.00 | $3,971.00 |
| G6. Payments made for monthly plans for technical services such as data plans and phone plans for HCW | $589.50 | $4.81 | $143.48 |
| **Per patient cost for Systems, Data management and technical support** | **$17.06** | **$8.11** | **$59.60** |
| h) Escalation in case of non-adherence and HCW response | | | |
| Escalation related phone calls: | | | |
| H1. Number of patients who required follow up by phone as part of adherence escalation procedure | 17 | 31 | 736 |
| H2. Average number of phone calls made by HCW per patient (for entire duration of the project) | 10 | 1 | 9.92 |
| H3. HCW time per call (in minutes) required to speak to a patient who requires adherence follow up | 10 | 2 | 1 |

| Cost components | Bangladesh | The Philippines | Tanzania |
|---|---|---|---|
| H4. Total time (in minutes) spent by HCW with patients on a phone for adherence follow up | 1,700 | 62 | 7,304 |
| H5. Type of HCW who usually makes phone calls | Research | TB nurse | Nurse |
| H6. Average HCW wage (per hour) | $1.36 | $1.59 | $1.09 |
| H7. Additional fees associated with phone call follow up (per call) | $0.00 | - | - |
| H8. Total cost for Phone calls | $38.42 | $1.64 | $132.69 |
| Escalation related Home Visits: | | | |
| H9. Number of patients who required a home visit as part of escalation procedure | 40 | 95 | 495 |
| H10.Average number of home visits per individual who requires home visits | 0 | 2 | 4 |
| H11.Amount of time per visit (including travel) | 180 | 45 | 120 |
| H12. Total time (in minutes) spent by HCW on home visits for patients who required adherence follow up | - | 8,550 | 237,600 |
| H13.Type of HCW doing home visit | Research | Volunteer | Volunteers |
| H14.HCW wage (per hour) | $1.36 | - | - |
| H15.Additional costs for HCW per visit (e.g., Travel/transport costs, incentives, etc.) | $1.77 | $2.30 | $1.79 |
| H16. Total cost for home visits | - | $437.00 | $3,544.20 |
| **Pro-rated per patient cost for escalation activity** | **$0.05** | **$1.11** | **$3.77** |
| i) DAT Training for HCWs *(Trainee costs)* | | | |
| 1st Category of HCW trained | Research | TB nurse | Doctors |
| I1.1 Number of Category 1 trained | 7 | 5 | 29 |
| I1.2 Typical amount of time for training (in minutes) (per HCW in category 1) | 1,440 | 300 | 600 |
| I1.3 Total time in minutes for training all HCW in category 1 | 10,080 | 1,500 | 17,400 |
| I1.4 HCW wage (per hour) | $1.36 | $1.59 | $2.40 |
| I1.5 Total cost for training all HCW in category 1 | $227.78 | $39.75 | $696.00 |
| 2nd Category of HCW trained | volunteer | Midwife | Clinicians |
| I2.1 Number of Category 2 trained | 0 | 2 | 16 |
| I2.2 Typical amount of time for training (in minutes) (per HCW in category 2) | 0 | 300 | 600 |
| I2.3 Total time in minutes for training all HCW in category 2 | - | 600 | 9,600 |
| I2.4 HCW wage (per hour) (category 2) | - | $1.20 | $1.09 |
| I2.5 Total cost for training all HCW in category 2 | - | $720.00 | $10,464 |
| 3rd Category of HCW trained | Research | TB Doctor | Nurses |
| I3.1 Number of Category 3 trained | 1 | 3 | 56 |
| I3.2 Typical amount of time for training (in minutes) (per HCW in category 3) | 1,440 | 300 | 600 |
| I3.3 Total time in minutes for training all HCW in category 3 | 1,440 | 900 | 33,600 |
| I3.4 HCW wage (per hour) (category 3) | $3.12 | $3.27 | $1.09 |
| I3.5 Total cost for training all HCW in category 3 | $4,499.06 | $2,943.00 | $36,624 |
| I4. Total cost for training | $4,726.85 | $3,702.75 | $47,784 |
| **Pro rated per patient training cost** | **$6.57** | **$9.35** | **$48.96** |
| j) Additional training costs *(Trainer costs)* | | | |
| Choose the training scenario for your project | Scenario I | | Scenario II |
| Scenario I: training was provided by individuals who were paid for each training session | | | |
| J1. Most common job category of staff providing the training sessions for HCW on how to use the 99DOTS | Research | IT | - |
| J2. Typical amount of time spent by a trainer on training (including preparation and delivery) (in hours) | 40 | 240 | - |
| J3. Approximate hourly salary of someone doing training (trainer) | $25.35 | $6.00 | - |
| J4. Number of trainers | 6 | 3 | - |
| J5. Total trainers' wage directly related to 99DOTS training | $6,083.64 | $4,320.00 | - |
| Scenario II: training was conducted by an outside organization or paid as a package | | | |
| J6. Total trainers' cost | - | $144.00 | - |
| J7. Additional expenditures related to training (including travel, training venue, subsistence for trainers and trainees, etc. but not time spent by, or salary paid to trainers or trainees)? Specify in column C and enter the cost in column C | - | $788.00 | $29,409.94 |
| **Pro rated per patient cost of running training sessions (not including time for HCW who are being trained)** | **$8.46** | **$2.35** | **$30.13** |
| **Total per patient cost for 99DOTS** | **$98.41** | **$105.97** | **$173.70** |

N=Numeric,
* Text in blue Indicates calculated fields
- indicates non applicable components

### Table s4. Cost components for VOT across the four study populations

| Cost components | Haiti | Moldova (All) | Moldova (DS-TB) | Moldova (DR-TB) | Philippines |
|---|---|---|---|---|---|
| Population | | | | | |
| I. Type of patients (DS-TB or DR-TB) | DS-TB | All | DS-TB | DR-TB | DR-TB |
| II. Length of a full course of treatment (in months) | 6 | 6 & 9 | 6 | 9 | 9 |
| III. Number of patients who were supported using VOT during the project | 87 | 308 | 173 | 135 | 110 |
| a) Phone and accessories | | | | | |
| A1. Number of phones that were purchased | 40 | 200 | 92 | 108 | 131 |
| A2. Unit cost of phone purchased | $69.0 | $110.0 | $110.0 | $110.0 | $97.7 |
| A3. Number of SIM cards that were purchased | 60 | 140 | 65 | 75 | 111 |
| A4. Unit cost of SIM card | $2.0 | - | - | - | $0.8 |
| A5. Number of charger or solar batteries that were purchased | 0 | - | - | - | 90 |
| A6. Unit cost of charger or solar batteries | $0.0 | - | - | - | $1.8 |
| A7. Number of phone lines provided with airtime | 60 | 384 | 177 | 207 | 54 |
| A8. Total cost for airtime per phone line for the entire duration of the project | $118.0 | $27.8 | $27.8 | $27.8 | $47.8 |
| A9. Total cost of phones (includes phone, SIM card and airtime) | $9,960.0 | $32,663.7 | $15,048.8 | $17,614.9 | $15,635.7 |
| **Pro rated per patient cost for phone and accessories** | **$114.5** | **$106.05** | **$86.99** | **$130.48** | **$142.1** |
| b) Data | | | | | |
| B1. Average size of a video recording in Megabytes (MB) | 60 | 30 | 30 | 30 | - |
| B2. Cost per MB of data | $0.0 | $0.0 | $0.0 | $0.0 | - |
| B3. Total number of video calls/recordings made by all patients in the project | 12,194 | 43,027 | 19,823 | 23,204 | 29,700 |
| B4. Total cost for data for all patients | $731.6 | $1,837.3 | $846.46 | $990.80 | $0.0 |
| **Per patient cost for data** | **$8.4** | **$5.97** | **$4.89** | **$7.34** | **$0.0** |
| c) Platform / infrastructure costs | | | | | |
| C1. Total amount spent on software licence (if applicable) for the entire duration of the project | $31,149.0 | - | - | - | $27,326.7 |
| C2. Initial Configuration costs (setting up VDOT platform in country) | - | - | - | - | - |
| C3. Total amount spent to develop a VDOT software or platform for a project not using an already existing system | - | $40,000.0 | $18,428.8 | $21,571.2 | - |
| **Per patient fixed cost of the platform/infrastructure** | **$358.0** | **$129.87** | **$106.52** | **$159.79** | **$248.4** |
| d) Adherence monitoring by HCW using VDOT platform | | | | | |
| Routine adherence monitoring using VDOT | | | | | |
| D1. Type of HCW who does adherence monitoring using VDOT platform | Field staff | Assistant | Assistant | Assistant | Nurse |
| D2. HCW wage (per hour) | $2.7 | $11.0 | $11.0 | $11.0 | - |
| D3. Typical amount of time (in min) spent by HCW monitoring adherence on the platform per patient (duration of project) | 701 | 540 | 360 | 540 | - |
| D4. Per patient cost of VDOT routine adherence monitoring | $31.5 | $99.00 | $66.00 | $99.00 | $0.0 |
| Video recordings viewing | | | | | |
| D5. Type of HCW who usually views video recordings | Officer | Assistant | Assistant | Assistant | Nurse |
| D6. HCW wage (per hour) | $1.7 | $11.0 | $11.0 | $11.0 | $2.9 |
| D7. Average number of minutes per patient per recording | 0 | 2 | 2 | 2 | 10 |
| D8. Average number of recordings made by a patient in the project | $140.2 | $139.70 | $114.59 | $171.88 | $270.0 |
| D9. Per patient average amount of time (in minutes) of recordings over the course of treatment | $42.0 | $279 | $229 | $171.88 | $2,700.0 |
| D10. Per patient cost of viewing video recordings | $1.2 | - | - | - | $132.7 |
| **Per patient cost of VDOT adherence monitoring (routine + viewing of recordings)** | **$32.7** | **$99.00** | **$66.00** | **$99.00** | **$132.7** |
| e) Systems, Data management and technical support | | | | | |
| E1. Amount spent for staff working on data management for VDOT platform (salaries paid for support of DAT platform) | $34,200.0 | - | - | - | - |
| E2. Number of days that technical support was provided or number of times specific technical services were provided (beyond initial setup) | 220 | 160 | 74 | 86 | - |
| E3. Cost per day or per activity reported in E2 | $55.0 | $28.5 | $28.5 | $28.5 | - |
| E4. Total cost of technical support for VDOT platform | $12,100.0 | $4,560.0 | $2,100.88 | $2,459.12 | $0.0 |
| E5. Amount paid for health facility hardware (laptops, desktop or tablet computer to track adherence) | $7,500.0 | $650.0 | $299.47 | $350.53 | $2,257.2 |
| E6. Payments made for monthly plans for technical services such as data plans and phone plans for HCW and health facility | $0.0 | $1,920.0 | $884.58 | $1,035.42 | - |

| Cost components | Haiti | Moldova (All) | Moldova (DS-TB) | Moldova (DR-TB) | Philippines |
|---|---|---|---|---|---|
| **Per patient cost for Systems, Data management and technical support** | **$618.4** | **$23.15** | **$18.99** | **$28.48** | **$20.5** |
| f) Escalation in case of non-adherence and HCW response | | | | | |
| Escalation related phone calls: | | | | | |
| F1. Number of patients who required follow up by phone as part of adherence escalation procedure | - | 32 | 15 | 17 | 42 |
| F2. Average number of phone calls made by HCW per patient (for entire duration of the project) | - | 21 | 21 | 21 | 50 |
| F3. Average HCW time per call (in minutes) required to speak to a patient who requires adherence follow up | - | 5 | 5 | 5 | 15 |
| F4. Total time ( in minutes) spent by HCW with patients on a phone for adherence follow up | - | 3,360 | 1,548 | 1,812 | 31,500 |
| F5. Type of HCW who usually makes phone calls | | Assistant | Assistant | Assistant | Nurse |
| F6. Average HCW wage (per hour) | - | $11.00 | $11.00 | $11.00 | $2.95 |
| F7. Additional fees associated with phone call follow up (per call) | - | - | - | - | - |
| F8. Total cost for Phone calls | $0.0 | $616.0 | $284.8 | $332.20 | $1,549 |
| Escalation related Home Visits: | | | | | |
| F9. Number of patients who required a home visit as part of escalation procedure | - | 0 | 0 | 0 | 18 |
| F10.Average number of home visits per individual who requires home visits | - | 0 | 0 | 0 | 6 |
| F11.Amount of time per visit (including travel) | - | 0 | 0 | 0 | 240 |
| F12. Total time ( in minutes) spent by HCW on home visits for patients who required adherence follow up | - | - | - | - | 25,920 |
| F13.Type of HCW doing home visit | - | Assistant | Assistant | Assistant | - |
| F14.HCW wage (per hour) | | $11.0 | $11.0 | $11.0 | $3.2 |
| F15.Additional costs for HCW per visit (e.g. Travel/transport costs, incentives, etc.) | | | | | |
| F16. Total cost for Home visits | $0.0 | $0.0 | $0.0 | $0.0 | $1,385.1 |
| **Pro rated per patient cost for escalation activity** | **$8.6** | **$2.00** | **$1.64** | **$2.46** | **$26.7** |
| g) DAT Training for HCWs (Trainee costs) | | | | | |
| 1st Category of HCW trained | | Doctors | Doctors | Doctors | Nurse |
| G1.1 Number of HCW trained | - | 43 | 43 | 43 | 6 |
| G1.2 Typical amount of time for training (in minutes) (per HCW in category 1) | - | 300 | 300 | 300 | 120 |
| G1.3 Total time in minutes for training all HCW in category 1 | - | 12,900 | 12,900 | 12,900 | 720 |
| G1.4 HCW wage (per hour) | - | $15.0 | $15.0 | $15.0 | $2.9 |
| G1.5 Total cost for training all HCW in category 1 | $0.0 | $3,225 | $3,225 | $3,225 | $35.4 |
| 2nd Category of HCW trained | | Assistant | Assistant | Assistant | Staff |
| G2.1 Number of HCW trained | - | 53 | 53 | 53 | 8 |
| G2.2 Typical amount of time for training (in minutes) (per HCW in category 2) | - | 420 | 420 | 420 | 240 |
| G2.3 Total time in minutes for training all HCW in category 2 | - | 22,260 | 22,260 | 22,260 | 1,920 |
| G2.4 HCW wage (per hour) (category 2) | - | $11.0 | $11.0 | $11.0 | - |
| G2.5 Total cost for training all HCW in category 2 | $0.0 | $4,081 | $4,081 | $4,081 | $0.0 |
| 3rd Category of HCW trained | - | N/A | N/A | N/A | Nurse |
| G3.1 Number of HCW trained | - | - | - | - | 4 |
| G3.2 Typical amount of time for training (in minutes) (per HCW in category 3) | - | - | - | - | 240 |
| G3.3 Total time in minutes for training all HCW in category 3 | - | - | - | - | 960 |
| G3.4 HCW wage (per hour) (category 3) | - | - | - | - | $2.9 |
| G3.5 Total cost for training all HCW in category 3 | $0.0 | $0.0 | $0.0 | $0.0 | $2,832 |
| G4. Total cost for training | $0.0 | $7,306 | $3,366 | $3,940 | $2,867 |
| **Pro rated per patient training cost** | **$13.8** | **$23.72** | **$19.46** | **$24.93** | **$26.1** |
| h) Additional training costs (Trainer costs) | | | | | |
| Choose the training scenario for your project | | | | | |
| Scenario I: training was provided by individuals who were paid for each training session | | | | | |
| H1. Most common job category of staff providing the training sessions for HCW on how to use the DAT | - | - | - | - | Doctor |
| H2. Typical amount of time spent by a trainer on training (including preparation and delivery) (in hours) | - | - | - | - | 8 |
| H3. Approximate hourly salary of someone doing training (trainer) | - | - | - | - | $30.8 |
| H4. Number of trainers | - | - | - | - | 1 |

| Cost components | Haiti | Moldova (All) | Moldova (DS-TB) | Moldova (DR-TB) | Philippines |
|---|---|---|---|---|---|
| H5. Total trainers' wage directly related to VDOT training | $0.0 | $0.0 | $0.0 | $0.0 | $246.2 |
| Scenario II: training was conducted by an outside organization or paid as a package | | | | | |
| H6. Total trainers' cost | - | - | - | - | - |
| H7. Additional expenditures related to training (including travel, training venue, subsistence for trainers and trainees, etc. but not time spent by or salary paid to trainers or trainees)? Specify in column C and enter the cost in column C | - | - | - | - | $7,034.62 |
| **Pro rated per patient cost of running training sessions (not including time for HCW who are being trained)** | **$0.0** | **$0.0** | **$0.0** | **$0.0** | **$64.0** |
| **Total per patient cost for VDOT is SUM of the following categories** | **$1,154.4** | **$389.76** | **$304.49** | **$452.48** | **$660.5** |

N=Numeric,
* Text in blue Indicates calculated fields
- indicates non applicable components

#### Table S5. Effectiveness

| Country | DAT | Study description | Effectiveness of DAT arm | Reference |
|---|---|---|---|---|
| Bangladesh | 99DOTS | Implementation study of 99DOTS in private sector TB screening and treatment centres established by icddr,b under its social enterprise model in Dhaka. | Overall adherence: 96% of prescribed doses taken | [1] |
| Philippines | 99DOTS | The study was designed to assess 99DOTS use in the private sector in the Philippines, where data suggests 50% of patients in the country seek care. | Overall adherence: 94% of prescribed doses taken # | [2] |
| United Republic of Tanzania | 99DOTS | The study was done in mining communities in Tanzania in four districts and four regions. The intervention involves (i) provision of medication in 99DOTS sleeves, (ii) delivery of reminders via SMS to patients, (iii) dosing histories used for counselling and for differentiated care (more intensive patient management), and (iv) targeted educational messaging based on adherence and risk factors via SMS or IVR. | Measured concordance with urine testing 95% of doses reported using 99DOTS were confirmed by urine testing . This concordance was highest (98%) in patients who were in their first 2 months of treatment. § | [3] |
| Haiti | VOT | A feasibility, acceptance, persistence, accuracy and sustainability study of VOT for prisoners in a low-income country | Median adherence of 85.7% of doses taken, but limited to the 65 patients who completed treatment | [4] |
| Philippines | VOT | This pilot study aimed to determine feasibility and acceptability of VOT in a high-burden, resource constrained DR-TB clinic in the Philippines where smartphones penetration is moderate and growing. | Good adherence was defined as ingestion of >90% of prescribed doses.<br>Observed doses:<br>**Males**<br>VOT 86.2%<br>Status quo 80.7%<br>**Females**<br>VOT 86.8%<br>Status quo 81.4% | [5] |
| Republic of Moldova | VOT | A pilot study to scale up locally developed VOT technology/program | Overall: 89% adherence to TB treatment *<br>DS-TB 92%<br>DR-TB 85% | [6] |

### Adherence Using 99DOTS calls as a proxy for adherence resulted in a 94% sensitivity using a urine test for isoniazid metabolites (IsoScreen) as the reference standard.
§ Similar urinalysis approach used in Tanzania to that in the Philippines to assess adherence
DAT; Digital adherence technology. DR-TB; drug resistant TB. DS-TB; drug susceptible TB. Icddr,b; International Centre for Diarrhoeal Disease Research, Bangladesh. KNCV; Koninklijke Nederlandse Centrale Vereniging tot bestrijding der Tuberculose. TB; Tuberculosis. VOT; video-observed treatment
* Adherence was measured by the proportion of days that a person with TB was observed ingesting medication during the planned treatment period

#### Table S6: DOT schedules and duration in the study settings – from the costing tool

| | 99DOTS sites | | | VOT sites | | | |
|---|---|---|---|---|---|---|---|
| | **Bangladesh** | **Philippines** | **Tanzania** | **Haiti** | **Moldova DS-TB** | **Moldova DR-TB** | **Philippines** |
| **Crude estimate for DOT cost for the standard of care - derived from the costing tool** | | | | | | | |
| Duration of TB treatment (in months) | 6 | 6 | 6 | 6 | 6 | 9 | 9 |
| Frequency of nurse visits (for clinical purposes) | 3 | 6 | 16 | 6 | 6 | 9 | 9 |
| Frequency of physician visits (for clinical purposes) | 2 | 6 | 6 | 6 | 6 | 9 | 36 |
| Proportion on DOT | 100% | 100% | 100% | 100% | 100% | 100% | 50% |
| Number of days DOT | 120 | 168 | 120 | 120 | 120 | 180 | 180 |
| Location of DOT - during the study | Health facility | Health facility | Home based | Prison | Health facility | Health facility | Health facility or Home |
| Hourly wage of person offering DOT support | $0.83 | $1.40 | $0.00 | $4.60 | $11.20 | $11.20 | $2.88 |
| Duration of DOT visit - in minutes | 45 | 45 | 0 | 45 | 15 | 15 | 2 |
